# Foundation versus Domain-Specific Models for Left Ventricular Segmentation on Cardiac Ultrasound

**DOI:** 10.1101/2023.09.19.23295772

**Authors:** Chieh-Ju Chao, Yunqi Richard Gu, Wasan Kumar, Tiange Xiang, Lalith Appari, Justin Wu, Juan M. Farina, Rachael Wraith, Jiwoon Jeong, Reza Arsanjani, Garvan C. Kane, Jae K. Oh, Curtis P. Langlotz, Imon Banerjee, Li Fei-Fei, Ehsan Adeli

**Author notes:** Li Fei-Fei and Ehsan Adeli are co-corresponding authors of this manuscript., **Corresponding Author:** Ehsan Adeli, Ph.D., Assistant Professor, Department of Psychiatry and Behavioral Sciences, Address: MC#5590, 1070 Arastradero Rd., Palo Alto, CA 94304.

## Abstract

The Segment Anything Model (SAM) was fine-tuned on the EchoNet-Dynamic dataset and evaluated on external transthoracic echocardiography (TTE) and Point-of-Care Ultrasound (POCUS) datasets from CAMUS (University Hospital of St Etienne) and Mayo Clinic (99 patients: 58 TTE, 41 POCUS). Fine-tuned SAM was superior or comparable to MedSAM. The fine-tuned SAM also outperformed EchoNet and U-Net models, demonstrating strong generalization, especially on apical 2-chamber (A2C) images (fine-tuned SAM vs. EchoNet: CAMUS-A2C: DSC 0.891 ± 0.040 vs. 0.752 ± 0.196, p<0.0001) and POCUS (DSC 0.857 ± 0.047 vs. 0.667 ± 0.279, p<0.0001). Additionally, SAM-enhanced workflow reduced annotation time by 50% (11.6 ± 4.5 sec vs. 5.7 ± 1.7 sec, p<0.0001) while maintaining segmentation quality. We demonstrated an effective strategy for fine-tuning a vision foundation model for enhancing clinical workflow efficiency and supporting human-AI collaboration.

## Introduction

Echocardiography provides a comprehensive anatomy and physiology assessment of the heart and is one of the most widely available imaging modalities in the field of Cardiology given its non-radiative, safe, and low-cost nature^1–3^. Cardiac chamber quantification, especially the left ventricle (LV), is one of the fundamental tasks of echocardiography studies in the current practice^4^, and the results can have direct effects on clinical decisions such as the management of heart failure, valvular heart diseases, and chemotherapy-induced cardiomyopathy ^5–8^. Although LV chamber quantification tasks are performed by trained sonographers or physicians, it is known to be subject to intra- and inter-observer variance, which can be up to 7%-13% across studies^9–12^. Many artificial intelligence (AI) applications have been applied to address this essential task and to minimize variations^3,13–18^.

While established AI systems are potential solutions, the training of segmentation AI models requires large amounts of training data and their corresponding expert-defined annotations, making them challenging and costly to implement^19^. In recent years, transformers, a type of neural network architecture with the self-attention mechanism that enables the model to efficiently capture complex dependencies and relationships, have revolutionized the field from natural language processing to computer vision^20–22^.

Vision transformers (ViT) ^20^ are a type of transformer specifically designed for images, which have shown impressive performance with simple image patches and have become a popular choice for the building of foundation models that can be fine-tuned for various applications^23^. Building on this success, Meta AI introduced the “Segment Anything Model” (SAM), a foundation large vision model that was trained on diverse datasets and that can adapt to specific tasks. This model achieves “zero-shot” segmentation, i.e., segments user-specified objects at different data resources without needing any training data^24^.

However, while the zero-shot performance of SAM on natural image datasets has been promising^24^, its performance on complex image datasets, such as medical images, has not been fully investigated. While not specifically including echocardiography images, one study tested SAM on different medical image datasets, including ultrasound, and the zero-shot performance was not optimal^25^. Recently, MedSAM was introduced as a universal tool for medical image segmentation. However, ultrasound or echocardiography was less represented in the training set despite > 1 million medical images being used^26^. This prompted a core research question: How can we effectively develop a segmentation model for a specific medical image segmentation task? Is it necessary to build a universal medical image segmentation model, or can a generic foundation model be fine-tuned with a relatively small, task-specific dataset to achieve comparable performance? In this context, we aim to study a novel strategy of using representative echocardiography examples for the fine-tuning of a foundation segmentation model (SAM) and compare its effectiveness with MedSAM and a state-of-the-art segmentation model trained with a domain-specific dataset (EchoNet)^3^.

We hypothesized that fine-tuning the foundation segmentation model with the above strategy can achieve similar or superior performance on LV segmentation compared to MedSAM or EchoNet. We further designed a human annotation study on a subset of images to investigate whether the model can enhance the efficiency of LV segmentation. Within the echocardiography domain, images obtained from different institutions and modalities (with different image qualities) were used to evaluate the generalization capability of SAM performance on external evaluation sets.

## Results

### SAM’s zero-shot performance on echocardiography and POCUS

We first tested the zero-shot performance of SAM. Overall, the zero-shot performance on the EchoNet-dynamic test set had a mean DSC of 0.863 ± 0.053. In terms of individual cardiac phase performance, end-diastolic frames were better than end-systolic frames (mean DSC 0.878 ± 0.040 vs. 0.849 ± 0.060) (**Table 1**). **Supplementary Table 1** summarizes the zero-shot performance on the EchoNet-dynamic training and validation sets. On the Mayo Clinic dataset, the mean DSC was 0.882 ± 0.036 and 0.861 ± 0.043 for TTE and POCUS, respectively. On the CAMUS dataset, we observed a mean DSC of 0.866 ± 0.039 and 0.852 ± 0.048 on A4C and A2C views, respectively (**Table 1**). When compared to the ground truth LVEF, the calculated LVEF had an MAE of 11.67%, 6.28%, and 6.38%, on EchoNet, Mayo-TTE, and Mayo-POCUS data, respectively (**Supplementary Table 2**).

**Table 1.** Comparison of Model Performance Based on Dice Similarity Coefficient (DSC) and Intersection over Union (IoU)

| Dataset | Phase | DSC |  |  |  |  | IoU |  |  |  |  |
| --- | --- | --- | --- | --- | --- | --- | --- | --- | --- | --- | --- |
|  |  | SAM<sub>112</sub><br>(fine-tuned) | SAM<br>(zero-shot) | MedSAM | EchoNet | U-Net | SAM<sub>112</sub><br>(fine-tuned) | SAM<br>(zero-shot) | MedSAM | EchoNet | U-Net |
| EchoNet-test | Overall | $0.911 \pm 0.045^{*\dagger\ddagger}$ | $0.863 \pm 0.053$ | $0.872 \pm 0.056$ | $0.915 \pm 0.047$ | $0.640 \pm 0.151$ | $0.840 \pm 0.071^{*\dagger\ddagger}$ | $0.763 \pm 0.077$ | $0.777 \pm 0.083$ | $0.847 \pm 0.072$ | $0.488 \pm 0.157$ |
| | ED | $0.929 \pm 0.030^{*\dagger\ddagger}$ | $0.878 \pm 0.040$ | $0.893 \pm 0.040$ | $0.903 \pm 0.052$ | $0.675 \pm 0.134$ | $0.868 \pm 0.050^{*\dagger\ddagger}$ | $0.784 \pm 0.062$ | $0.809 \pm 0.062$ | $0.826 \pm 0.078$ | $0.524 \pm 0.148$ |
| | ES | $0.894 \pm 0.050^{*\dagger\ddagger}$ | $0.849 \pm 0.060$ | $0.850 \pm 0.061$ | $0.928 \pm 0.038$ | $0.606 \pm 0.160$ | $0.812 \pm 0.077^{*\dagger\ddagger}$ | $0.742 \pm 0.084$ | $0.744 \pm 0.088$ | $0.868 \pm 0.059$ | $0.453 \pm 0.159$ |
| Mayo-TTE | Overall | $0.902 \pm 0.032^{*\dagger\ddagger}$ | $0.882 \pm 0.036$ | $0.883 \pm 0.039$ | $0.893 \pm 0.090$ | $0.597 \pm 0.169$ | $0.822 \pm 0.051^{*\dagger\ddagger}$ | $0.790 \pm 0.056$ | $0.792 \pm 0.061$ | $0.814 \pm 0.093$ | $0.444 \pm 0.162$ |
| | ED | $0.916 \pm 0.024^{*\dagger\ddagger}$ | $0.889 \pm 0.037$ | $0.903 \pm 0.027$ | $0.916 \pm 0.031$ | $0.634 \pm 0.162$ | $0.846 \pm 0.040^{*\dagger\ddagger}$ | $0.802 \pm 0.058$ | $0.825 \pm 0.044$ | $0.846 \pm 0.051$ | $0.482 \pm 0.161$ |
| | ES | $0.887 \pm 0.032^{*\dagger\ddagger}$ | $0.875 \pm 0.033$ | $0.862 \pm 0.039$ | $0.870 \pm 0.119$ | $0.561 \pm 0.169$ | $0.799 \pm 0.050^{*\dagger\ddagger}$ | $0.779 \pm 0.052$ | $0.760 \pm 0.059$ | $0.782 \pm 0.113$ | $0.408 \pm 0.156$ |
| Mayo-POCUS | Overall | $0.857 \pm 0.047^{++}$ | $0.861 \pm 0.043$ | $0.846 \pm 0.072$ | $0.667 \pm 0.279$ | $0.396 \pm 0.239$ | $0.753 \pm 0.070^{++}$ | $0.758 \pm 0.066$ | $0.740 \pm 0.102$ | $0.554 \pm 0.265$ | $0.275 \pm 0.192$ |
| | ED | $0.878 \pm 0.036^{++}$ | $0.876 \pm 0.032$ | $0.868 \pm 0.064$ | $0.717 \pm 0.255$ | $0.463 \pm 0.232$ | $0.785 \pm 0.056^{++}$ | $0.781 \pm 0.051$ | $0.772 \pm 0.092$ | $0.607 \pm 0.253$ | $0.329 \pm 0.194$ |
| | ES | $0.836 \pm 0.047^{++}$ | $0.846 \pm 0.048$ | $0.825 \pm 0.074$ | $0.617 \pm 0.295$ | $0.330 \pm 0.229$ | $0.720 \pm 0.069^{++}$ | $0.735 \pm 0.072$ | $0.708 \pm 0.103$ | $0.501 \pm 0.270$ | $0.221 \pm 0.175$ |
| CAMUS-A2C | Overall | $0.891 \pm 0.040^{*\dagger\ddagger}$ | $0.852 \pm 0.048$ | $0.857 \pm 0.057$ | $0.752 \pm 0.196$ | $0.460 \pm 0.154$ | $0.805 \pm 0.062^{*\dagger\ddagger}$ | $0.745 \pm 0.069$ | $0.754 \pm 0.081$ | $0.633 \pm 0.196$ | $0.311 \pm 0.130$ |
| | ED | $0.897 \pm 0.037^{*\dagger\ddagger}$ | $0.860 \pm 0.042$ | $0.877 \pm 0.040$ | $0.754 \pm 0.196$ | $0.458 \pm 0.135$ | $0.815 \pm 0.059^{*\dagger\ddagger}$ | $0.756 \pm 0.062$ | $0.783 \pm 0.060$ | $0.635 \pm 0.197$ | $0.307 \pm 0.116$ |
| | ES | $0.885 \pm 0.041^{*\dagger\ddagger}$ | $0.845 \pm 0.052$ | $0.838 \pm 0.065$ | $0.751 \pm 0.196$ | $0.461 \pm 0.172$ | $0.795 \pm 0.064^{*\dagger\ddagger}$ | $0.734 \pm 0.073$ | $0.725 \pm 0.088$ | $0.632 \pm 0.196$ | $0.315 \pm 0.144$ |
| CAMUS-A4C | Overall | $0.897 \pm 0.036^{*\dagger\ddagger}$ | $0.866 \pm 0.039$ | $0.877 \pm 0.044$ | $0.850 \pm 0.097$ | $0.418 \pm 0.159$ | $0.815 \pm 0.058^{*\dagger\ddagger}$ | $0.766 \pm 0.059$ | $0.784 \pm 0.066$ | $0.749 \pm 0.117$ | $0.277 \pm 0.128$ |
| | ED | $0.904 \pm 0.033^{*\dagger\ddagger}$ | $0.873 \pm 0.037$ | $0.890 \pm 0.035$ | $0.850 \pm 0.098$ | $0.428 \pm 0.139$ | $0.827 \pm 0.054^{*\dagger\ddagger}$ | $0.776 \pm 0.056$ | $0.804 \pm 0.055$ | $0.749 \pm 0.119$ | $0.282 \pm 0.114$ |
| | ES | $0.889 \pm 0.038^{*\dagger\ddagger}$ | $0.860 \pm 0.041$ | $0.864 \pm 0.048$ | $0.850 \pm 0.096$ | $0.409 \pm 0.176$ | $0.803 \pm 0.060^{*\dagger\ddagger}$ | $0.756 \pm 0.061$ | $0.763 \pm 0.070$ | $0.749 \pm 0.115$ | $0.272 \pm 0.140$ |

\*p < 0.05, Zero-shot vs. fine-tuned SAM. †p < 0.05, Fine-tuned SAM vs. EchoNet model. ‡p < 0.05, Fine-tuned SAM vs. MedSAM. ++p < 0.05, Fine-tuned SAM vs. U-Net. A2C: apical 2 chamber view, A4C: apical 4 chamber view, CAMUS: Cardiac Acquisitions for Multi-structure Ultrasound Segmentation, DSC: Dice Similarity Score, ED: end-diastolic, ES: end-systolic, IoU: Intersection over Union, SAM: segment anything model, TTE: transthoracic echocardiography, POCUS: point-of-care ultrasound. Data expressed as mean±standard deviation.

**Table 2.** Comparison of Model Performance Based on Hausdorff Distance (HD) and Average Surface Distance (ASD)

| Dataset | Phase | HD | ASD |
| --- | --- | --- | --- |

|  |  | SAM<sub>112</sub><br>(fine-tuned) | SAM<br>(zero-shot) | MedSAM | EchoNet | U-Net | SAM<sub>112</sub><br>(fine-tuned) | SAM<br>(zero-shot) | MedSAM | EchoNet | U-Net |
| --- | --- | --- | --- | --- | --- | --- | --- | --- | --- | --- | --- |
| Echo<br>Net-<br>test | Overall | 4.408 ± 1.999<sup>†‡</sup> | 7.780 ± 3.155 | 6.381 ± 2.143 | 9.196 ± 2.929 | 13.596 ± 6.131 | 0.156 ± 0.133<sup>†‡</sup> | 0.350 ± 0.211 | 0.286 ± 0.181 | 0.753 ± 0.371 | 1.785 ± 1.429 |
|  | ED | 4.425 ± 2.066<sup>†‡</sup> | 8.320 ± 3.274 | 6.490 ± 2.272 | 9.715 ± 2.966 | 14.060 ± 6.637 | 0.123 ± 0.098<sup>†‡</sup> | 0.338 ± 0.202 | 0.240 ± 0.150 | 0.860 ± 0.395 | 1.668 ± 1.340 |
|  | ES | 4.390 ± 1.931<sup>†‡</sup> | 7.239 ± 2.935 | 6.271 ± 2.000 | 8.678 ± 2.799 | 13.132 ± 5.543 | 0.188 ± 0.154<sup>†‡</sup> | 0.361 ± 0.219 | 0.331 ± 0.198 | 0.646 ± 0.309 | 1.903 ± 1.505 |
| Mayo-TTE | Overall | 7.543 ± 4.780<sup>†‡</sup> | 8.581 ± 4.495 | 4.435 ± 1.275<sup>§</sup> | 23.714 ± 6.585 | 13.896 ± 5.261 | 0.814 ± 0.930<sup>†‡</sup> | 0.854 ± 0.928 | 0.401 ± 0.147<sup>§</sup> | 5.705 ± 2.984 | 1.877 ± 1.425 |
|  | ED | 7.855 ± 5.225<sup>†‡</sup> | 9.127 ± 4.847 | 4.920 ± 1.402<sup>§</sup> | 26.539 ± 6.527 | 13.067 ± 4.669 | 0.903 ± 1.061<sup>†‡</sup> | 0.927 ± 1.011 | 0.412 ± 0.165<sup>§</sup> | 5.337 ± 2.502 | 1.585 ± 1.214 |
|  | ES | 7.241 ± 4.333<sup>†‡</sup> | 8.054 ± 4.101 | 3.951 ± 0.914<sup>§</sup> | 20.938 ± 5.389 | 14.697 ± 5.701 | 0.728 ± 0.783<sup>†‡</sup> | 0.785 ± 0.843 | 0.390 ± 0.126<sup>§</sup> | 6.066 ± 3.375 | 2.159 ± 1.562 |
| Mayo-POCUS | Overall | 8.804 ± 6.716<sup>†‡</sup> | 9.072 ± 6.291 | 3.666 ± 1.041<sup>§</sup> | 19.821 ± 5.776 | 24.452 ± 23.324 | 1.202 ± 1.311<sup>†‡</sup> | 1.172 ± 1.297 | 0.436 ± 0.234<sup>§</sup> | 5.421 ± 2.655 | 4.911 ± 5.545 |
|  | ED | 8.273 ± 6.395<sup>†‡</sup> | 9.033 ± 5.851 | 3.858 ± 1.035<sup>§</sup> | 20.059 ± 5.617 | 20.855 ± 19.624 | 0.972 ± 1.113<sup>†‡</sup> | 1.065 ± 1.248 | 0.362 ± 0.150<sup>§</sup> | 4.889 ± 2.061 | 3.792 ± 4.414 |
|  | ES | 9.335 ± 7.061<sup>†‡</sup> | 9.111 ± 6.776 | 3.475 ± 1.023<sup>§</sup> | 19.595 ± 5.983 | 28.050 ± 26.268 | 1.431 ± 1.462<sup>†‡</sup> | 1.279 ± 1.351 | 0.511 ± 0.278<sup>§</sup> | 5.927 ± 3.058 | 6.029 ± 6.341 |
| CAM<br>US-A2C | Overall | 5.624 ± 2.121<sup>†‡</sup> | 5.842 ± 2.081 | 5.169 ± 2.354<sup>§</sup> | 6.962 ± 3.951 | 18.709 ± 7.665 | 0.237 ± 0.147<sup>†‡</sup> | 0.257 ± 0.154 | 0.220 ± 0.214<sup>§</sup> | 0.469 ± 0.482 | 3.277 ± 2.000 |
|  | ED | 6.120 ± 2.198<sup>†‡</sup> | 6.302 ± 2.071 | 5.294 ± 2.365<sup>§</sup> | 6.653 ± 3.676 | 20.610 ± 8.075 | 0.236 ± 0.155<sup>†‡</sup> | 0.252 ± 0.131 | 0.193 ± 0.155<sup>§</sup> | 0.384 ± 0.294 | 3.412 ± 1.907 |
|  | ES | 5.129 ± 1.920<sup>†‡</sup> | 5.382 ± 1.990 | 5.043 ± 2.339<sup>§</sup> | 7.270 ± 4.190 | 16.808 ± 6.720 | 0.237 ± 0.139<sup>†‡</sup> | 0.262 ± 0.173 | 0.248 ± 0.257<sup>§</sup> | 0.555 ± 0.603 | 3.142 ± 2.081 |
| CAM<br>US-A4C | Overall | 5.001 ± 1.728<sup>†‡</sup> | 5.601 ± 1.926 | 4.826 ± 2.515<sup>§</sup> | 5.763 ± 2.519 | 19.748 ± 7.868 | 0.205 ± 0.117<sup>†‡</sup> | 0.230 ± 0.124 | 0.176 ± 0.173<sup>§</sup> | 0.370 ± 0.267 | 3.717 ± 1.938 |
|  | ED | 5.367 ± 1.850<sup>†‡</sup> | 6.085 ± 1.919 | 5.223 ± 2.895<sup>§</sup> | 5.838 ± 2.523 | 21.822 ± 8.322 | 6.120 ± 2.198<sup>†</sup> | 6.302 ± 2.071 | 5.294 ± 2.365<sup>§</sup> | 6.653 ± 3.676 | 20.610 ± 8.075 |
|  | ES | 4.635 ± 1.512<sup>†‡</sup> | 5.118 ± 1.809 | 4.428 ± 1.990<sup>§</sup> | 5.687 ± 2.515 | 17.674 ± 6.787 | 5.129 ± 1.920<sup>†‡</sup> | 5.382 ± 1.990 | 5.043 ± 2.339<sup>§</sup> | 7.270 ± 4.190 | 16.808 ± 6.720 |

\*p < 0.05, Zero-shot vs. fine-tuned SAM. †p < 0.05, Fine-tuned SAM vs. EchoNet model. ‡p < 0.05, Fine-tuned SAM vs. MedSAM. §p < 0.05, Fine-tuned SAM vs. U-Net. A2C: apical 2 chamber view, A4C: apical 4 chamber view, CAMUS: Cardiac Acquisitions for Multi-structure Ultrasound Segmentation, ASD: Average Surface Distance, HD: Hausdorff Distance, ED: end-diastolic, ES: end-systolic, IoU: Intersection over Union, SAM: segment anything model, TTE: transthoracic echocardiography, POCUS: point-of-care ultrasound. Data expressed as mean ± standard deviation.

**Table 3.**
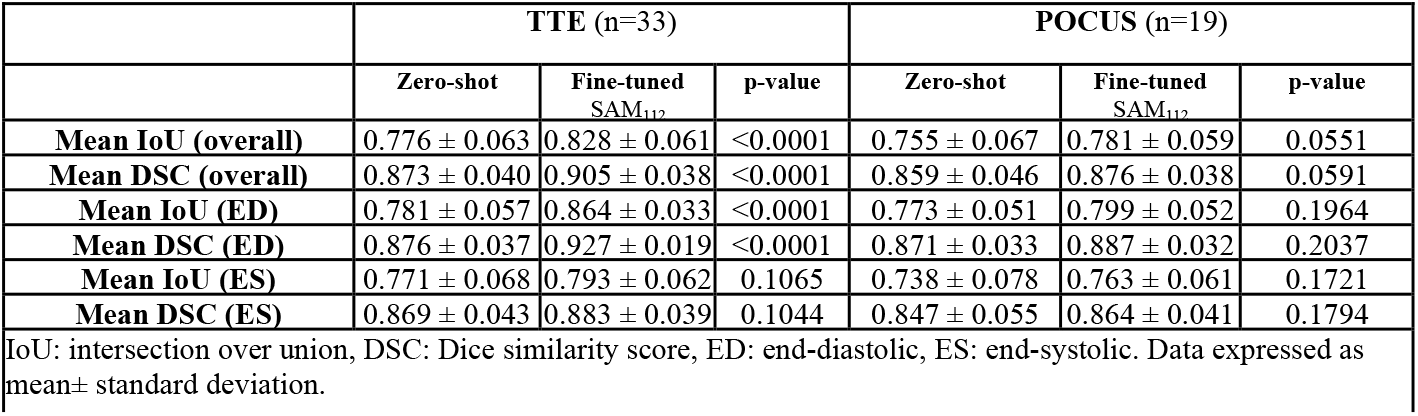
Zero-shot vs. Fine-tuned SAM_112_ performance on TTE and POCUS (against the second observer).

### SAM’s fine-tuned performance on echocardiography and POCUS

Compared to zero-shot, fine-tuning generally improved the performance of SAM, with a mean DSC of 0.911 ± 0.045 (SAM_112_) on the EchoNet-dynamic test set (**Table 1** and **Supplementary Table 1**). Similar improvement was also observed in Mayo TTE data, with an overall mean DSC of 0.902 ± 0.032. In contrast, no significant improvement was observed on the POCUS data (DSC 0.857 ± 0.047), while the performance was numerically improved when compared with the ground truth of the second observer (DSC 0.876 ± 0.038), as summarized in **Table 3**. The EchoNet model had a significant performance drop, especially on the Mayo-POCUS and CAMUS A2C datasets (**Table 1**). When compared to the ground truth LVEF, the calculated LVEF had an MAE of 7.52%, 5.47%, and 6.70%, on the EchoNet test, Mayo-TTE, and Mayo-POCUS data, respectively (**Supplementary Table 2**).

Qualitatively, poor-performing cases often featured suboptimal images (such as weak LV endocardial borders, off-axis views; **Figure 1**, Panels c, f), highlighting the importance of image quality (**Figure 1**). On a representative POCUS case, fine-tuned SAM_112_ predicted masks that were more consistent with LV geometry (**Figure 2**).

**Figure 1.**
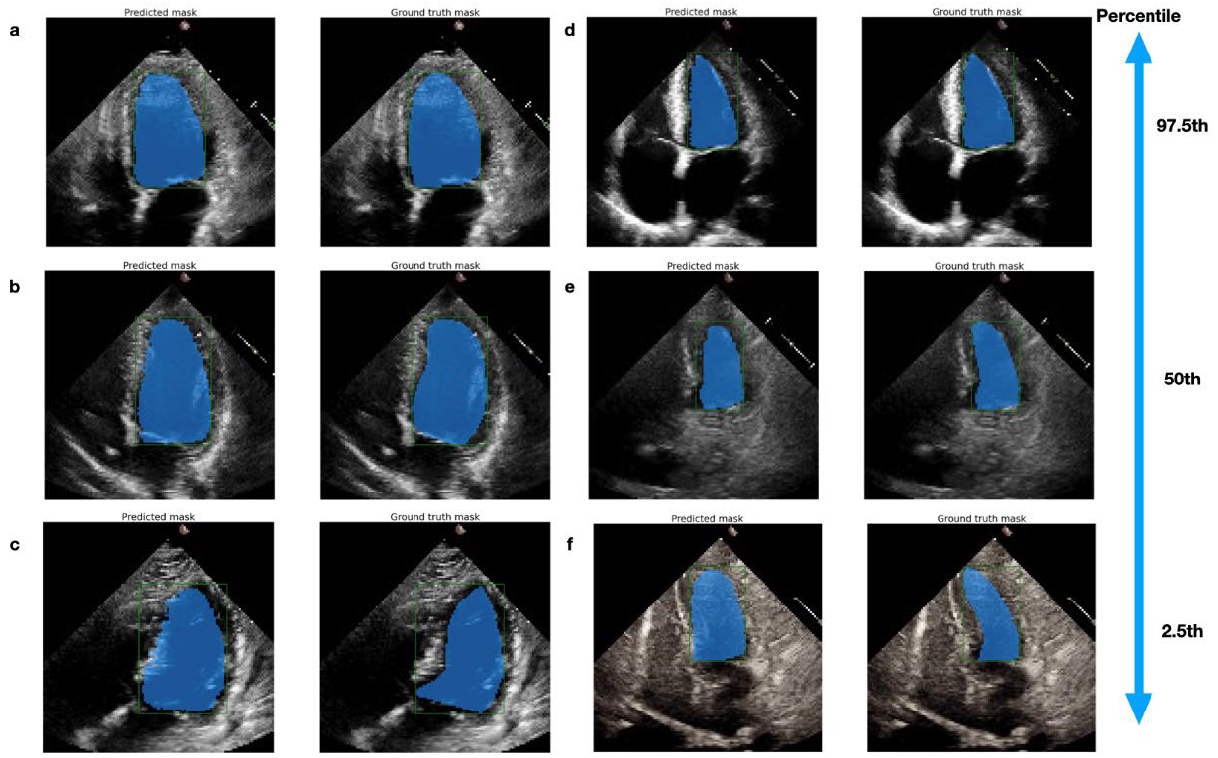
Qualitative performance of fine-tuned SAM on representative cases against ground truth on the EchoNet-dynamic test dataset. From top to bottom: 97.5th to 2.5th percentile of DSC. Panels a, b, and c are end-diastolic frames, and Panels d, e, and f are end-systolic frames. We observed that many of the poor-performance cases had suboptimal image qualities, such as weak LV endocardial borders or off-axis views (Panels c and f), suggesting the importance of good input image quality on model performance. Additionally, end-diastolic frames usually have a better delineation of borders than end-systolic frames, which is consistent with the model performance (end-diastolic slightly better than end-systolic).

**Figure 2.**
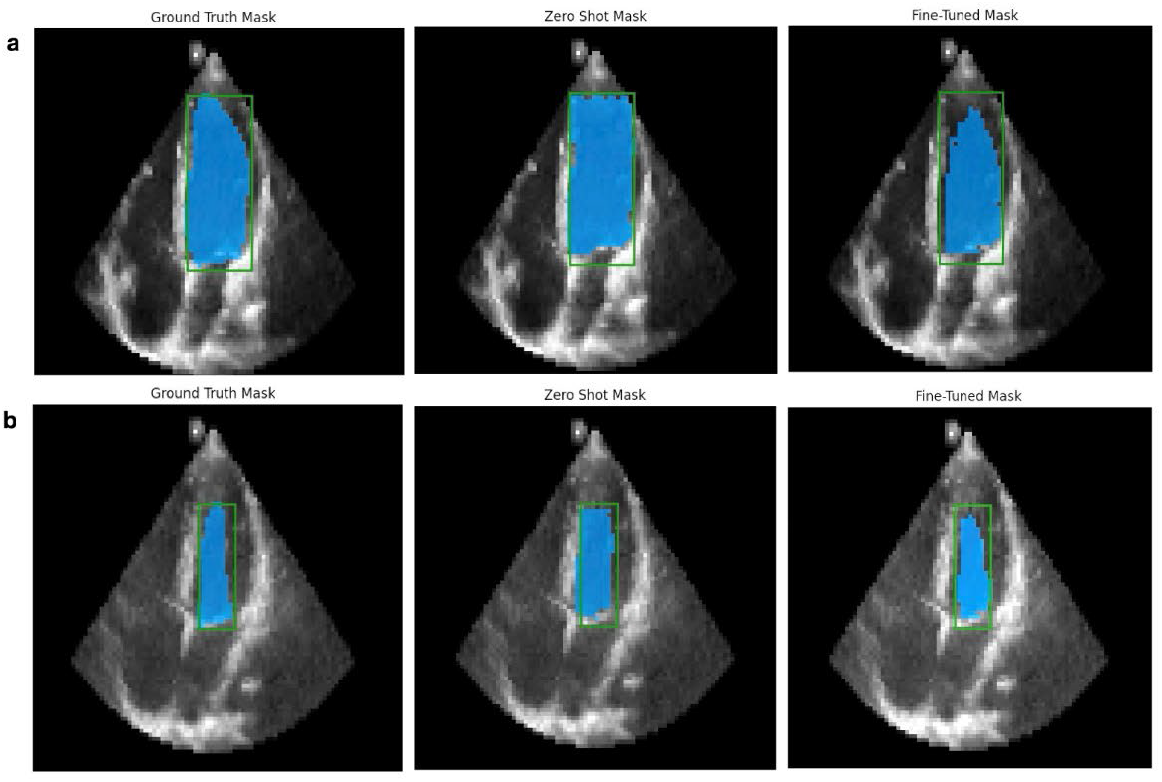
Zero-shot and fine-tuned SAM performance on a representative POCUS case. **Panel a**. end-diastolic frame, **Panel b**. end-systolic frame. From left to right are the ground truth, zero-shot, and fine-tuned mask, with an overlay of bounding boxes (green-colored) and mask (blue-colored), on the original POCUS image. Fine-tuned masks were more consistent with the anticipated left ventricular geometry on visualization. Note that POCUS images generally had worse quality compared to transthoracic echocardiography images.

### Comparison of SAM, MedSAM, EchoNet, and U-Net models

Comparisons were made between fine-tuned SAM_112_ (fine-tuned using 112× 112 images), MedSAM, EchoNet, and U-Net. Under the same setting, the inference time for SAM was about 7.3 images/sec, MedSAM was 5.0 images/sec, EchoNet was 153.3 images/sec, and UNet was 26 images/sec. Note that EchoNet takes video input, so the inference time was averaged by the number of frames in each video.

When evaluated by DSC and IoU, EchoNet demonstrated a similar level of performance compared to fine-tuned SAM_112_ on the EchoNet test set (SAM_112_ vs. EchoNet: DSC 0.911 ± 0.045 vs. 0.915 ± 0.047, p<0.0001) and the Mayo TTE dataset (DSC 0.902 ± 0.032 vs. 0.893 ± 0.090, p<0.0001). However, EchoNet significantly underperformed on the Mayo POCUS and CAMUS datasets, with a performance drop ranging from 5-25% (**Table 1**). MedSAM also showed about 2-5% worse performance than fine-tuned SAM_112_across all datasets (all p<0.0001), except for the CAMUS dataset, as it was part of MedSAM’s training data. U-Net had worse performance than fine-tuned SAM_112_ across all datasets (all p< 0.0001) (**Table 1**). When evaluated by HD and ASD, MedSAM had the best performance across most of the datasets, while fine-tuned SAM_112_ had the best performance on the EchoNet test set. This was followed by SAM (zero-shot), EchoNet, and then U-Net (**Supplementary Table 3**).

Further fine-tuned SAM_1024_ (fine-tuned using 1024× 1024 images as MedSAM) essentially achieved the same performance as fine-tuned MedSAM, with a DSC of 0.935 and 0.936, respectively. Both of the fine-tuned models also demonstrated similar performance across different datasets, except for the CAMUS dataset, which is part of MedSAM’s training set (**Table 4**). We also observed that after the finetuning, both SAM_1024_ and MedSAM had dropped performance slightly on the Mayo and POCUS datasets, compared to fine-tuned SAM_112_ and base MedSAM.

**Table 4.**
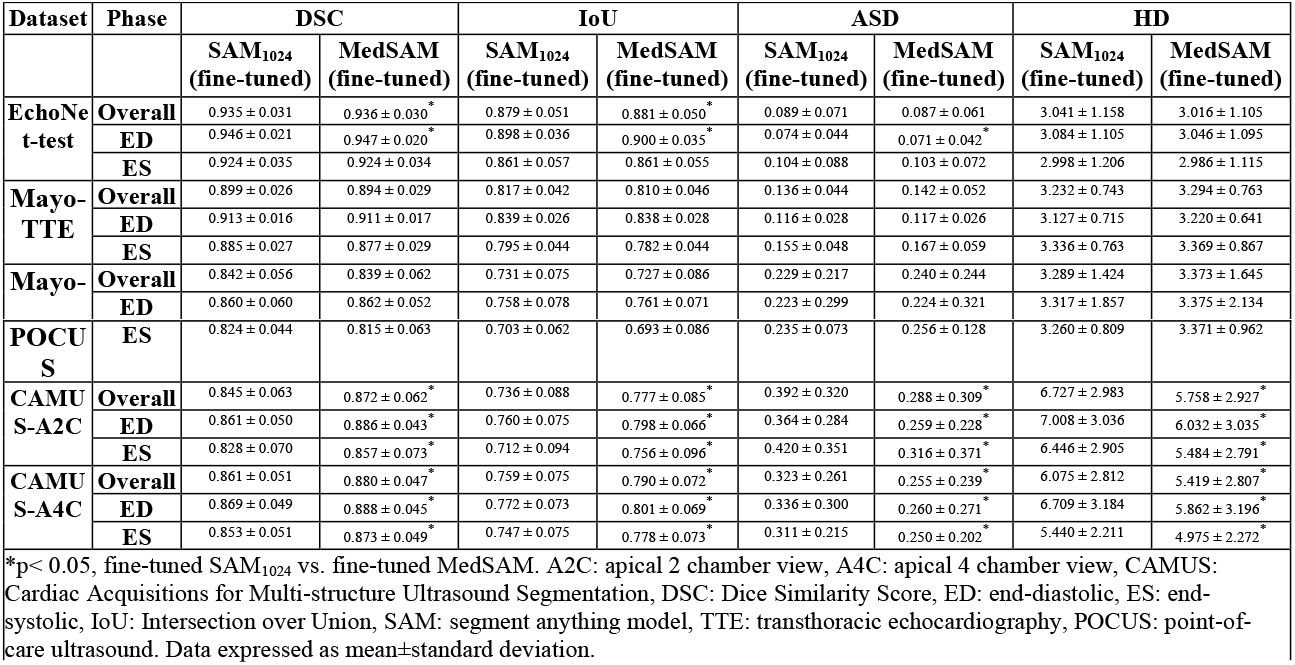
Comparison of fine-tuned SAM and MedSAM.

### Human Annotation Study

To test SAM’s potential in enhancing clinical workflow, we designed a human annotation study using SAM_112_. In the study, we observed that AI assistance significantly improved efficiency by decreasing 50% annotation time (p <0.0001; **Figure 3a**), which remains true for both expert-level and medical student-level annotators (**Figure 3b and 3c**). The AI-assisted workflow maintained the quality of left ventricle segmentation, as measured by DSC (**Figure 3d, e, and f**). Qualitative assessments indicated an improvement in performance for inexperienced annotators, which may not be completely captured by the DSC scores (**Supplementary Figure 1**).

**Figure 3.**
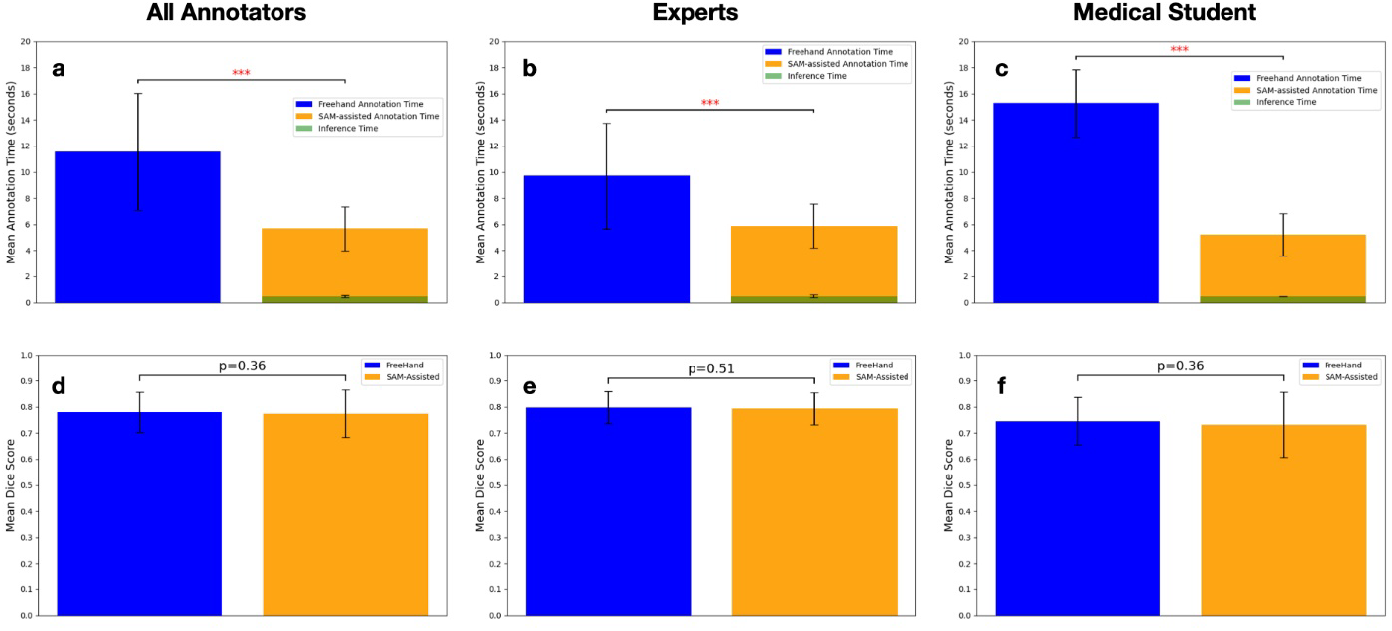
SAM-assisted workflow decreased the time for annotation while maintaining the quality of echocardiography image segmentation. Panel a. With SAM’s assistance, the average annotation time significantly decreased by 50%. Panel b. Among experienced annotators, the workflow decreased annotation time by 39.2%; Panel c. It also reduced 66.0% annotation time for an inexperienced annotator. Panel d, e, and f. No significant difference was observed in segmentation quality when measured by the Dice Similarity Score. ***p<0.0001.

## Discussion

The major contributions of this work include: 1) presenting a data-efficient and cost-effective strategy for training a LV segmentation model for echocardiography images based on SAM and leveraging its generalization capability for POCUS images, and 2) showing the potential of optimizing clinical workflows through a human-in-the-loop approach, where SAM significantly improved efficiency while maintaining annotation quality.

Echocardiography, like other ultrasound modalities, is generally considered an imaging modality with more challenges due to its operator dependency and low signal-to-noise ratio^3,27–29^. Additionally, objects could often have weak border linings or be obstructed by artifacts on ultrasound/echocardiography images, which posed specific challenges for echocardiography segmentation tasks^3,25^. While the image/frame-based approach of SAM does not consider consecutive inter-frame changes and spatio-temporal features of the echocardiogram, fine-tuned SAM (both SAM_112_ and SAM_1024_) achieved superior frame-level segmentation performance on all datasets compared to the video-based EchoNet model^3^. Importantly, there were cases with suboptimal image quality and imperfect human labels (**Figure 1**) in the EchoNet-dynamic data set^3,30^, which can limit the model performance.

In terms of generalization capability, the fine-tuned SAM_112_ model demonstrated robust performance on unseen TTE (including the A2C view in CAMUS) and POCUS data, with about a 1% and 5% drop in performance, respectively. In contrast, there was a significant drop in EchoNet and UNet’s performance on the POCUS dataset (**Table 1**). Similarly, when evaluated by border-sensitive metrics such as HD and ASD, foundation models (SAM and MedSAM) were able to generate more consistent borders compared to EchoNet and U-Net (**Supplementary Table 3**). This again demonstrated the advantage of leveraging the generalization capabilities of foundation models ^23^ in building AI solutions for the rapidly growing use of POCUS in cardiac imaging.^34^ While evaluation on a larger POCUS dataset is required to better evaluate inter-observer variations, we demonstrated a strategy to fine-tune foundation models using readily available and relatively high-quality TTE results, knowing that POCUS images usually come with larger variations in operator skill levels, image quality, and scanning modalities^34–37^.

On a head-to-head comparison, fine-tuned SAM_112_ outperformed base MedSAM across almost all datasets when evaluated by DSC and IoU (**Table 1)**. This is likely due to the fact that cardiac ultrasound images were less represented in MedSAM’s training set of over 1 million images. When fine-tuned under the same setting, SAM_1024_ and fine-tuned MedSAM can achieve the same level of performance and generalization capability on a specialized LV segmentation task (**Table 4**). Our study supports a data-efficient foundation model training strategy that uses representative examples for specific tasks in medical subspecialties, providing a generalizable solution that overcomes the challenge of large-scale, high-quality dataset collection in this field^19,31^. Importantly, starting with SAM also saved computational resources for training on 20 A100 (80G) GPUs in MedSAM’s training process^26^.

SAM’s interactive capability allows the potential integration of SAM into research or clinical workflows in echocardiography labs to create high-quality segmentation masks at scales ^23,25^. We demonstrated the potential of a SAM-based, human-in-loop workflow for LV segmentation, significantly reducing the annotation time and maintaining the segmentation quality across users with different experience levels (**Figure 3; Supplementary Figure 1**). Compared to fully automatic models^3^, an interactive approach ensures that segmentations can be directly prompted or modified to the level of interpreters’ satisfaction^25,32^. This process also has the potential to gain the trust of human users to facilitate integration^33^. While future studies are needed to assess its real impact on clinical practice, the interactive functionality of SAM could be especially helpful in challenging cases or when fully automatic models fail to predict segmentations accurately ^25^.

This study is limited by its retrospective nature and could be subject to selection bias. Since SAM is an image-based model, its performance was evaluated on pre-selected end-diastolic and end-systolic frames rather than the beat-to-beat assessment of the video as proposed by the EchoNet-Dynamic model.

However, we still demonstrated superior frame-level performance with fine-tuned SAM. While adapters are another potential direction to use SAM for echocardiography segmentation^39^, we have not specifically explored this approach in the current paper. Since this manuscript focuses on exploring effective strategies for adapting a generic foundation model like SAM, we did not modify the architecture, and as such, the study does not include substantial technical innovations. The balance of performance and generalization capability was not comprehensively explored. Additionally, how SAM can be integrated into the current echocardiography lab workflow and its real-world effects will need to be validated in a prospective setting, which is beyond the scope of the study.

In conclusion, fine-tuning SAM on a relatively small yet representative echocardiography dataset resulted in superior performance compared to the state-of-the-art echocardiography segmentation model, EchoNet, as well as base MedSAM, and achieved the same performance as fine-tuned MedSAM. We demonstrated a data-efficient and cost-effective approach for handling specialized complex echocardiography images, and SAM’s potential to enhance current echocardiography workflows by improving the efficiency with a human-in-the-loop design.

## Methods

### Population and Data Curation

#### EchoNet-Dynamic

The EchoNet-Dynamic dataset is publicly available (https://echonet.github.io/dynamic/); details of the dataset have been described previously ^3^. In brief, the dataset contains 10,030 apical-4-chamber (A4C) TTE videos at Stanford Health Care in the period of 2016-2018. The raw videos were preprocessed to remove patient identifiers and downsampled by cubic interpolation into standardized 112 × 112-pixel videos. Videos were randomly split into 7,465, 1,277, and 1,288 patients, respectively, for the training, validation, and test sets^3^. The mean left ventricular ejection fraction of the EchoNet-dynamic dataset was 55.8±12.4%, 55.8±12.3%, and 55.5±12.2%, for the training, validation, and test set, respectively^3^. Patient characteristics were not available for this public dataset. In this study, the cases without ground truth labels were excluded from this analysis (5 from the train set, 1 from the test set) (**Supplementary Table 3**).

#### Mayo Clinic

A dataset from the Mayo Clinic (Rochester, MN) that includes 99 randomly selected patients (58 TTE in 2017-2018 and 41 point-of-care ultrasound studies (POCUS) in 2022) was used as an external validation dataset. The A4C videos were reviewed by a clinical sonographer (RW) and a cardiologist (JMF). Fifty-two (33 TTE and 19 POCUS) out of the total 99 cases were traced by both of the annotators to select and segment the end-diastolic and end-systolic frames. The tracings were done manually on commercially available software (MD.ai, Inc., NY). In the Mayo Clinic data set (n=99), the mean age was 47.5 ± 17.8 years, 58 (58.6%) were male, and coronary artery disease, hypertension, and diabetes were found in 20 (20.2%), 42 (42.4%), and 15 (15.2%) of patients, respectively. The dataset contains an A4C view of 58 TTE cases and 41 POCUS cases, the LVEF was 61.7 ± 7.5% for TTE and 63.2 ± 11.9% for POCUS cases.

#### CAMUS

The Cardiac Acquisitions for Multi-structure Ultrasound Segmentation (CAMUS) dataset contains 500 cases from the University Hospital of St Etienne (France) with detailed tracings on both A4C and A2C views ^40^. The CAMUS cohort (n=500) had a mean age of 65.1 ± 14.4 years, 66% male, and a mean LVEF of 44.4 ± 11.9%.

### Segment Anything Model (SAM), MedSAM, and U-Net

The Segment Anything Model (SAM) is an image segmentation foundation model trained on a dataset of 11 million images and 1.1 billion masks^32^. It can generate object masks from input prompts like points or boxes. SAM’s promptable design enables zero-shot transfer to new image distributions and tasks, achieving competitive or superior performance compared to fully supervised methods ^32^. In brief, the model comprises a VisionEncoder, PromptEncoder, MaskDecoder, and Neck module, which collectively process image embeddings, point embeddings, and contextualized masks to predict accurate segmentation masks ^32^. MedSAM is a fine-tuned version of the base SAM, trained on over 1.5 million medical image-mask pairs across 10 modalities and 30 cancer types, with trained weights available ^26^. *U-Net* architecture was selected as the representative of commonly used segmentation deep learning models. In our study, the U-Net architecture^41^ leveraged a ResNet-18 backbone^42^.

### Data Preprocessing

Each EchoNet-Dynamic video (112 × 112 pixels, in avi format) was exported into individual frames without further resizing. End-diastolic and end-systolic frames of each case were extracted, which corresponded to the human expert-traced, frame-level ground truth segmentation coordinates in the dataset. Ground truth segmentation masks were generated according to the coordinates and saved in the same size (112 × 112 pixels). The labeled frames of Mayo TTE and POCUS images were exported from the MD.ai platform and followed a similar preprocessing method to remove patient identifiers, then horizontally flipped and resized to 112 × 112 pixels^3^. The CAMUS images were rotated by 270 degrees and resized to be consistent with the EchoNet format. When importing to the SAM model, all the raw images were resized with the built-in function “ResizeLongestSide.”^32^

### Zero-shot Evaluation

The original SAM ViT-base model (model type “vit_b”, checkpoint “sam_vit_b_01ec64.pth “) without modification or fine-tuning was used to segment the dataset, referred to as zero-shot learning in machine learning literature^26,32^. The larger versions of SAM (ViT Large and ViT Huge) were not used as they did not offer significant performance improvement despite higher computational demands^32^. Bounding box coordinates of each left ventricle segmentation tracing were generated from the ground truth segmentations and used as the prompt for SAM^24^. Of note, while SAM supports both the bounding box and point prompts, our study focused exclusively on the bounding box prompt due to the incomplete and relatively weak borders in echocardiography images. Additionally, since the left atrium and LV are connected structures, point prompts may result in masks that incorrectly link the two chambers.

### Model Fine-tuning

The SAM ViT-base model was used for fine-tuning with a procedure described by Ma et al ^38^. We used the training set cases (n=7,460) of the EchoNet-Dynamic as our customized dataset without data augmentation. The same bounding box was used as the prompt, as described above. We used a customized loss function, which is the unweighted sum of Dice loss and cross-entropy loss^38,43^. Adam optimizer^44^ was used (weight decay = 0), with an initial learning rate of 2e-5 (decreased to 3e-6 over 27 epochs). The batch size was 8. The model was fine-tuned on a node on the Stanford AI Lab cluster with a 24 GB NVIDIA RTX TiTAN GPU. The fine-tuning procedure of U-Net followed the same procedure except for using bounding boxes, and the training epoch was 23 for the best-performing model. No data augmentation was performed.

To ensure fair comparison between SAM and MedSAM, we used MedSAM’s fine-tuning setup with 1024 × 1024 images, and both models were trained for 70 epochs. The two versions of fine-tuned SAM models were referred to as SAM_112_ and SAM_1024_, respectively. The training time for SAM_112_ and SAM_1024_ was about 25 hours and 40 hours, respectively. MedSAM required a training time of about 50 hours. UNet required 16 mins.

### Validation and Generalization

The fine-tuned SAM was tested on the test set of the EchoNet-Dynamic dataset. To test the generalization capacity of SAM, we used external validation samples from the CAMUS dataset (both A2C and A4C) ^40^ and a Mayo Clinic dataset including the A4C view of cases of TTE and POCUS devices.

### Human Annotation Study

We conducted a human annotation study to evaluate the enhancement of segmentation efficiency and quality when utilizing a fine-tuned SAM. Three annotators (two experts and one medical student) independently annotated 100 echocardiography images, randomly selected from the EchoNet-Dynamic test set. Each one annotated 100 images with and without SAM assistance. In the SAM-assisted workflow, annotators were instructed to draw minimal bounding boxes to prompt the model. In the regular workflow, annotators manually trace the endocardium contour, as per standard clinical practice. Annotators were allowed to refine their annotations until they were satisfactory. We recorded and compared the time required to complete annotations and the quality of annotations (DSC against ground truth) using both workflows.

### Statistical Model Performance Evaluation

The model segmentation performance was directly evaluated by Intersection over Union (IoU), Dice similarity coefficient (DSC), Hausdorff Distance (HD), and Average Surface Distance (ASD) against human ground truth labels^45^. IoU and DSC assess the overlap of segmented areas, while HD and ASD evaluate the alignment of segmentation borders. The formulas are provided in **Supplementary Note 1**. Depending on the normality of distribution, two-tailed, paired t-tests or Wilcoxon tests were conducted to assess the statistical significance of the differences in accuracy between models (zero-shot vs. fine-tuned SAM, EchoNet vs. fine-tuned SAM, and MedSAM vs. fine-tuned SAM), as well as the annotation time and quality metrics in the human annotation study (AI-assisted vs. non-AI-assisted), with p<0.05 as significant.

### Ethical review and approval

*All the studies have been performed in accordance with the Declaration of Helsinki*

EchoNet-Dynamic dataset contains a publicly available, de-identified dataset that was approved by Stanford University Institutional Review Board and data privacy review through a standardized workflow by the Center for Artificial Intelligence in Medicine and Imaging (AIMI) and the University Privacy Office. The CAMUS dataset is also publicly available, under the approval of the University Hospital of St Etienne (France) ethical committee after full anonymization. The use of the Mayo Clinic dataset was approved by the institutional review board (protocol#22-010944); only patients providing informed consent for minimal-risk retrospective studies were included, thus the requirement for additional informed consent was waived.

## Data Availability

The EchoNet and CAMUS datasets are publicly available. The Mayo Clinic dataset can not be made publicly available due to patient privacy regulations; it is available from the corresponding author upon reasonable request.

## Code Availability

We released a checkpoint of the fine-tuned SAM and MedSAM models, along with the fine-tuning, inference, and statistical analysis code. The code and checkpoint are available on GitHub: https://github.com/chiehjuchao/SAM-Echo.git

## Acknowledgment

We would like to acknowledge Owen Crystal, MS, for his contributions to the organization of EchoNet-related data, which significantly facilitated the analysis. The authors declare no funding support for this work.

## Author Contributions

C.J.C.: Conceptualization, methodology, software, validation, data curation, formal analysis, writing – original draft, visualization.

Y.R.G.: Methodology, software.

W.K.: Software, data curation.

T.X.: Methodology, software.

L.A.: Methodology, software.

J.W.: Methodology, software.

J.M.F.: Data curation.

R.W.: Data curation.

J.J.: Data curation, software.

R.A.: Data curation, resources, writing – review and editing.

G.C.K.: Data curation, resources, writing – review and editing.

J.K.O.: Data curation, resources, writing – review and editing.

C.P.L.: Resources, writing – review and editing.

I.B.: Conceptualization, resources, writing – review and editing, supervision.

F.F.L.: Conceptualization, methodology, resources, writing – review and editing, supervision.

E.A.: Conceptualization, methodology, resources, writing – review and editing, supervision.

All authors have read and approved the manuscript.

## Abbreviations

AI: Artificial intelligence
A2C: apical 2 chamber, echocardiography view
A4C: apical 4 chamber, echocardiography view
CAMUS: Cardiac Acquisitions for Multi-structure Ultrasound Segmentation
IoU: Intersection over the union
DSC: Dice similarity coefficient
LV: Left ventricle
POCUS: Point-of-care ultrasound
SAM: Segment anything model
TTE: Transthoracic echocardiography
ViT: Vision Transformer

