## Supplemental Table 1 for "Foundation versus Domain-Specific Models for Left Ventricular Segmentation on Cardiac Ultrasound"

**Supplementary Note 1. Formulas of evaluation metrics.**

1. Intersection over Union (IoU)

The IoU measures the overlap between the predicted segmentation and the ground truth segmentation.

$$IoU = \frac{|A \cap B|}{|A \cup B|}$$

Where:

- A is the set of pixels in the predicted segmentation.
- B is the set of pixels in the ground truth segmentation.
- $|A \cap B|$ is the number of overlapping pixels between AAA and BBB.
- $|A \cup B|$ is the total number of unique pixels in AAA and BBB.

2. Dice Similarity Coefficient (DSC)

The DSC is another overlap metric, emphasizing both precision and recall.

DSC=$\frac{2 \cdot |A \cap B|}{|A|+|B|}$

Where:

- A is the predicted segmentation set.
- B is the ground truth segmentation set.
- |A| and |B| are the total number of pixels in the respective segmentations.

3. Hausdorff Distance (HD)

The HD measures the greatest distance from a point in one boundary to the closest point in the other boundary.

$$HD(A, B) = \max\left(\sup_{a \in A} \inf_{b \in B} \|a - b\|, \sup_{b \in B} \inf_{a \in A} \|b - a\|\right)$$

Where:

- A and B are the boundaries of the predicted and ground truth segmentations, respectively.
- sup is the supremum (maximum).
- inf is the infimum (minimum).
- $\|a-b\|$ is the Euclidean distance between points a and b.

4. Average Surface Distance (ASD)

The ASD calculates the average distance between the boundary points of the predicted and ground truth segmentations.

$$\mathrm{ASD}(A, B) = \frac{1}{|A|} \sum_{a \in A} \min_{b \in B} \|a - b\|$$

Where:

- A and B are the boundaries of the predicted and ground truth segmentations, respectively.
- $|A|$ and $|B|$ are the number of boundary points in A and B.
- $\min\|a-b\|$ is the shortest distance from a point $a \in A$.
- The sum is taken over all points in set A, and the minimum distance to a point in set B is used.

**Supplementary Table 1. Zero-shot and Fine-tuned SAM performance on EchoNet-dynamic Train and Validation set**

| | **Zero-shot** | | **Fine-tuned** | |
|---|---|---|---|---|
| | **EchoNet-Dynamic-Train** | **EchoNet-Dynamic-Validation** | **EchoNet-Dynamic-Train** | **EchoNet-Dynamic-Validation** |
| **Mean IoU (overall)** | 0.761 ± 0.080 | 0.762 ± 0.078 | 0.842 ± 0.070 | 0.841 ± 0.069 |
| **Mean DSC (overall)** | 0.862 ± 0.055 | 0.862 ± 0.054 | 0.913 ± 0.044 | 0.912 ± 0.043 |
| **Mean IoU (ED)** | 0.781 ± 0.068 | 0.783 ± 0.065 | 0.868 ± 0.052 | 0.869 ± 0.050 |
| **Mean DSC (ED)** | 0.875 ± 0.046 | 0.877 ± 0.043 | 0.928 ± 0.032 | 0.929 ± 0.030 |
| **Mean IoU (ES)** | 0.740 ± 0.085 | 0.740 ± 0.084 | 0.816 ± 0.076 | 0.813 ± 0.074 |

| | | | | |
|---|---|---|---|---|
| **Mean DSC (ES)** | 0.848 ± 0.060 | 0.848 ± 0.059 | 0.897 ± 0.049 | 0.895 ± 0.047 |
| IoU: intersection over union, DSC: Dice similarity score, ED: end-diastolic, ES: end-systolic. Data expressed as mean± standard deviation. | | | | |

**Supplementary Table 2. LVEF prediction task evaluated by R-square and MAE of zero-shot vs. fine-tuned SAM**

| | **EchoNet-dynamic test** | | **CAMUS-A2C** | | **CAMUS-A4C** | | **Mayo-TTE** | | **Mayo-POCUS** | |
|---|---|---|---|---|---|---|---|---|---|---|
| | **Zero-shot** | **Fine-tuned** | **Zero-shot** | **Fine-tuned** | **Zero-shot** | **Fine-tuned** | **Zero-shot** | **Fine-tuned** | **Zero-shot** | **Fine-tuned** |
| **R-squared** | 0.141 | 0.161 | 0.445 | 0.761 | 0.549 | 0.709 | 0.374 | 0.517 | 0.638 | 0.718 |
| **MAE (%)** | 11.7 | 7.52 | 9.75 | 6.11 | 9.67 | 7.31 | 6.28 | 5.47 | 6.38 | 6.70 |
| The summation of disks method was used to calculate the LV ejection fraction (LVEF) from the end-diastolic and end-systolic segmentation masks. While the model was not trained for LVEF prediction, the model-derived LVEF measurements were compared to the ground truth-derived LVEF by R-square and mean absolute error (MAE). | | | | | | | | | | |

**Supplementary Table 3. Excluded EchoNet-dynamic dataset cases due to missing ground truth**

| **Filename** | **Split** |
|---|---|
| 0X234005774F4CB5CD.avi | TRAIN |
| 0X2DC68261CBCC04AE.avi | TRAIN |
| 0X35291BE9AB90FB89.avi | TRAIN |
| 0X6C435C1B417FDE8A.avi | TRAIN |
| 0X5515B0BD077BE68A.avi | TRAIN |
| 0X5DD5283AC43CCDD1.avi | TEST |

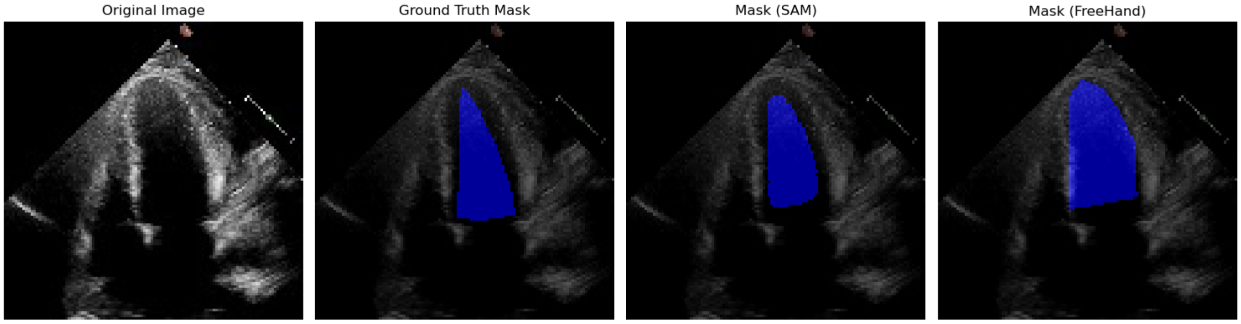


**Supplementary Figure 1.** This representative qualitative example illustrates the impact of SAM-assisted workflow on improving segmentation quality for inexperienced users. The panels, from left to right, display the original image, the ground truth mask overlay, the SAM-assisted mask overlay, and the freehand mask overlay. This example highlights an imperfect ground truth mask, showing that the SAM-assisted annotation is substantially better than the freehand annotation. Notably, the freehand annotation incorrectly included the septal and lateral wall, which is inappropriate for left ventricular segmentation.
